# Modelling the coronavirus disease (COVID-19) outbreak on the Diamond Princess ship using the public surveillance data from January 20 to February 20, 2020

**DOI:** 10.1101/2020.02.26.20028449

**Authors:** Shi Zhao, Peihua Cao, Daozhou Gao, Zian Zhuang, Marc KC Chong, Yongli Cai, Jinjun Ran, Kai Wang, Yijun Lou, Weiming Wang, Lin Yang, Daihai He, Maggie H Wang

## Abstract

The novel coronavirus disease 2019 (COVID-19) outbreak on the Diamond Princess ship has caused over 634 cases as of February 20, 2020. We model the transmission process on the ship with a stochastic model and estimate the basic reproduction number at 2.2 (95%CI: 2.1-2.4). We estimate a large dispersion parameter than other coronaviruses, which implies that the virus is difficult to go extinction. The epidemic doubling time is at 4.6 days (95%CI: 3.0-9.3), and thus timely actions were crucial. The lesson learnt on the ship is generally applicable in other settings.

## Backgrounds

The coronavirus disease 2019 (COVID-19), caused by the severe acute respiratory syndrome coronavirus 2 (SARS-CoV-2), emerged in Wuhan, China in the end of 2019, and spread to over 40 foreign countries in a short period of time [1-3]. The World Health Organization (WHO) declared the outbreak to be a public health emergency of international concern on January 30, 2020 [4]. To date, there are over 70000 COVID-19 confirmed cases in China, and over 2000 cases outside China. The COVID-19 outbreak on the Diamond Princess, a British cruise ship that contains 3711 tourists and crew members, caused 634 confirmed cases as of February 20, 2020 and became the largest outbreak outside China [5].

In this study, we model the COVID-19 outbreak on the Diamond Princess ship, estimate the key epidemiological parameters of this outbreak and explore the effects of several different control measures.

## Methods

We consider the population on the Diamond Princess ship as a close cohort with *N* = 3711 individuals. Following [6], we simulate the transmission process stochastically. For a given COVID-19 case with symptom onset on the *t*-th day, the number of secondary cases follows a negative binomial (NB) distribution with a mean at *R*_eff_(*t*) and a dispersion parameter at *k* [7]. The *R*_eff_(*t*) is the effective reproduction number, and *R*_eff_ (*t*) = *R*_0_[*N* – *C*(*t*)]/*N*, where *C*(*t*) is the cumulative number of cases at the *t*-th day and *R*_0_ is the basic reproduction number of COVID-19 to be estimated. The time delay between symptom onset dates of a primary case and any of its associated secondary cases is the serial interval (SI). Hence, for a given primary case with symptom onset on the *t*-th day, the symptom onset time of each associated secondary case is expected to be the summation of *t* and SI of COVID-19. It is conventional to assume the SI to follow a Gamma distribution with mean at 4.5 days and standard deviation (SD) at 3.1 days [8, 9].

We collect the confirmed cases time series released in the situation reports of COVID-19 infections in Japan [5]. We simulate the courses of the COVID-19 outbreaks stochastically starting from one infectious index case on January 20, 2020. We calculate the maximum likelihood estimates of *R*_0_ and *k* by fitting model to the number of confirmed cases with Poisson-distributed likelihood framework. The 95% confidence intervals (95%CI) of *R*_0_ and *k* are calculated by using the profile likelihood estimation framework with a cutoff threshold determined using a Chi-square quantile [10].

We repeat the fitting and estimation procedures above under an alternative scenario with mean SI at days and SD at 3.4 days, which was estimated in [2]. We evaluate the modelling performance with mean SI at 4.5 or 7.5 days by the Akaike information criterion (AIC). Under the later scenario with mean SI at 7.5 days, we find that the *R*_0_ is larger than 5 which is not in line with WHO estimates and hard to justify in a ‘quarantine’ setting. In particular, the later scenario yields a higher AIC by 10 units than the former scenario with mean SI at 4.5 days. Thus, we argue the former scenario is most likely the reality, and consider it as the main results.

Furthermore, we consider four ‘what-if’ scenarios and they were

- scenario (**0**): the best-fit model, i.e., baseline scenario;
- scenario (**1**): 1000 susceptibles were removed on February 11, 2020;
- scenario (**2**): the *R*_0_ was reduced to 1.5; and
- scenario (**3**): combining scenarios (**1**) and (**2**).

We simulate the model under four different scenarios and estimate the key epidemiological parameters. We estimate the cumulative number of cases on February 20, the outbreak final size, the epidemic doubling time and the peaking time to capture the patterns of outbreak.

## Results

With a mean SI at 4.5 days, we estimate the *R*_0_ to be 2.2 (95%CI: 2.1-2.4), which is consistent with previous estimates [7, 8, 11]. The fitting results are shown in Fig 1(a) and (b), which match the observed data well. We estimate the dispersion term (*k*) to be 44 (95%CI: 6-88), which is significantly larger than 1 and consistent with [7]. The simulation results with or without dispersion term (*k*) were largely consistent, which implies the virus is hard to go extinction that other two coronavirus. Note that low *k* implies high chance of superspreading events and high chance of extinction.

**Figure 1.**
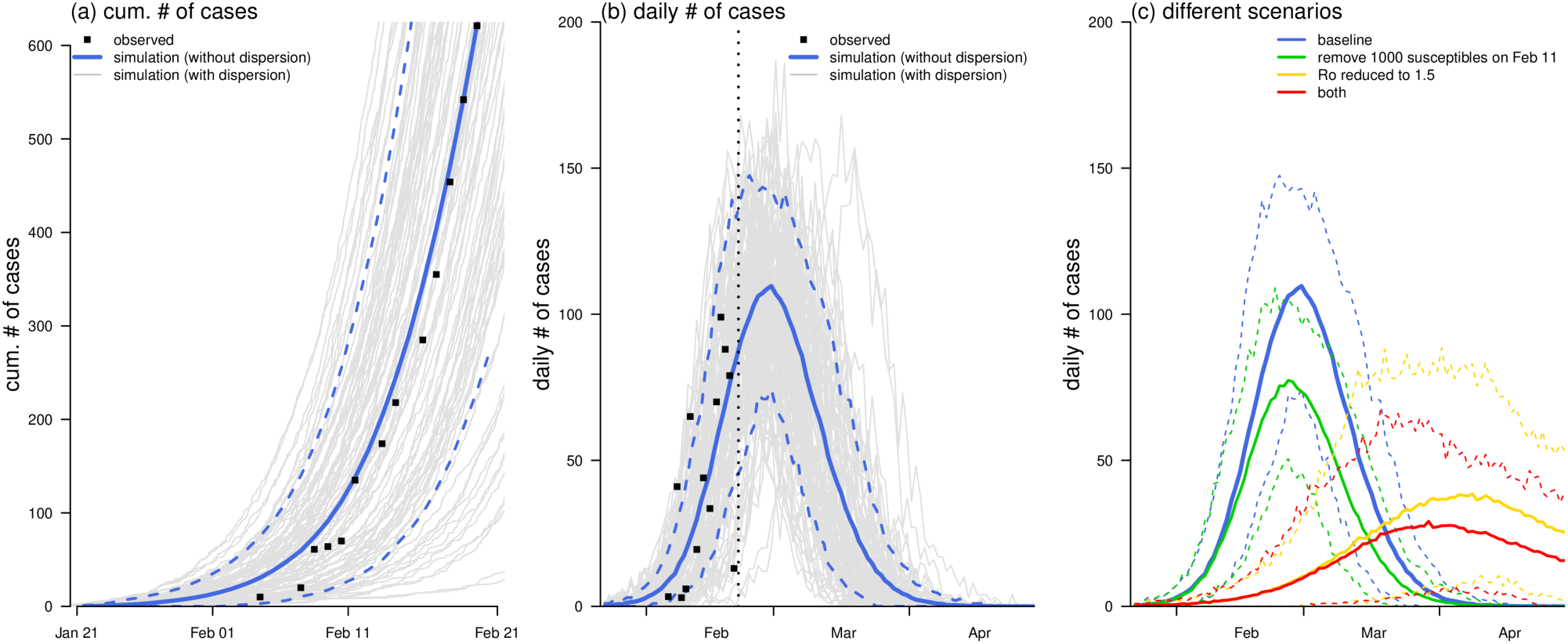
The observed (dots) and fitted (curves) number of COVID-19 cases on the Diamond Princess ship. Panel (a) shows the cumulative number of cases, and panel (b) shows the daily number of cases. In panels (a) and (b), the black dots are the observed number of cases time series, the blue curves are the simulation median (bold) and 95% CI (dashed), and the grey curves are 1000 simulation samples. In panel (b), the vertical black dashed line is February 20, 2020. Panel (c) shows the estimated epidemic curves under different scenarios. The bold curves are the simulation median, and the dashed curves are the 95% CIs. In panels (b) and (c), the blue curves show the baseline scenario, and they are the same.

Without control measures, we estimate that the COVID-19 outbreak is likely to cause 3066 cases (95%CI: 2046-3441) on the ship, and the epidemic curve is likely to peak around February 28, 2020 with a doubling time at 4.6 days (95%CI: 3.0-9.3), see Fig 1(b) and Table 1. Under scenario (**1**), timely reduction in the susceptible population could lower the final size and sustainably reduce the daily incidences. Under scenario (**2**), the lower the *R*_0_, decreasing from 2.2 to 1.5, the lower the final size and the later the peaking time will be. If the susceptible pools and *R*_0_ are reduced simultaneously, the COVID-19 outbreak on the ship will be likely mitigated and postponed largely, see Fig 1(d).

**Table 1.** Summary of the key epidemiological estimates of the outbreak under different scenarios.

| scenario | Description | cum. # on Feb 20 | final size | doubling time (day) | peaking time |
| --- | --- | --- | --- | --- | --- |
| (0) | Baseline | 657 (296, 1298) | 3066 (2046, 3441) | 4.6 (3.0, 9.3) | Feb 28 (Feb 20, Mar 6) |
| (1) | remove 1000 susceptibles on Feb 11 | 557 (248, 1076) | 2238 (1486, 2548) | 5.2 (3.1, 20.4) | Feb 26 (Feb 19, Mar 5) |
| (2) | $R_0$ reduced to 1.5 | 39 (10, 133) | 1702 (639, 3249) | 3.8 (1.7, 8.1) | Apr 8 (Mar 17, Apr 27) |
| (3) | both (1) and (2) | 39 (8, 138) | 1332 (481, 2506) | 3.1 (1.0, 10.7) | Apr 6 (Mar 13, Apr 27) |

## Discussion

The *R*_0_ of COVID-19 on the Diamond Princess ship is likely to be lower than that of the severe acute respiratory syndrome (SARS) which ranges from 2.2 to 3.6 [12]. The *k* quantifies the chance of the superspreading events and the extinction of the disease. We obtained a large *k* which implies that COVID-19 is much easier to persist than other coronavirus with a small *k* [6, 7, 13,14]. On the other hand, our finding highlights the different characteristics between COVID-19 (less superspreading) and SARS (more superspreading), which should be taken into consideration into mitigation. As such, the self-protection actions including wearing facemasks, reducing outdoor activities or gathering and maintain hygiene and sterilized, were thus recommended.

The situation on the Diamond Princess cruise implies that the virus could spread rapidly, most likely with a short SI than previously estimated [2]. Note that a recent study on the 1099 patients found that the median of incubation period is only 3 days, thus a short effective SI is also possible [15]. With a shorter SI, a relatively lower *R*_0_ could also result in a rapid growth of the epidemic size [16], and with a shorter epidemic doubling time as we show in Table 1. Therefore, timely contact tracing and effectively quarantine were crucial to shutoff the transmission.

The simulation results under scenarios (**1**)-(**3**) indicate that the public health control measures could effectively mitigate the COVID-19 outbreak on the ship in terms of the final size, see Table 1. Decreasing the disease transmissibility in terms of *R*_0_ could postpone the peak, which may gain valuable time to prepare and allocate the resources in response to incoming patients. Relocating the population at risk (if possible) could sustainably decrease the daily incidences, and thus relieve the tensive demands in the healthcare, and improve in the treatment outcome.

We suggest maintaining the enhancement in both populational level public health control as well as the individual level self-protection actions in combating the COVID-19 outbreak. With detailed information on public intervention, the analytical framework in this study can be extended to a complex context and used for evaluating the effects of certain control measures.

From Japan official reports (https://www.mhlw.go.jp/stf/newpage_09542.html), we obtain the following

2/15/2020 陽性が確認されたのは、延べ9 3 0 名の検査中2 8 5 名 (うち無症状病原体保有者延べ7 3 名) となりました。

2/20/2020 陽性が確認されたのは、延べ3 ,0 6 3 名の検査中 6 3 4 名(うち無症状病原体保有者延べ3 2 8 名) となりました。

English Translation

2/15/2020 Positive cases were confirmed in 285 of the 930 people tested (including 73 asymptomatic pathogen carriers).

2/20/2020 A total of 634 people (including 328 asymptomatic pathogen carriers) tested positive for a total of 3,063 people.

From the above official announcement of Japan, the cumulative ratio of asymptomatic cases out of all confirmations went up from 25.6% to 51.7% between Feb 15 and Feb 20, 2020. It is hard to believe that such a large amount of asymptomatic cases were infected before the quarantine start date (Feb 5, 2020) and missed in Feb 15 test. The number of tests are much larger than the number of positive results, and this implies high effort of testing and relatively reliable data. It is reasonable to argue that the quarantine is not so successful to prevent the emergence of asymptomatic cases, but successfully slowed down the occurrence of symptomatic cases, as suggested by studies based on cases onset date data. In the other words, quarantine measure likely changed the way how the virus transmitted, thus the level of symptoms of cases. Presumably, in these studies of onset date data only, asymptomatic confirmations are not counted as cases. In this study, we use laboratory confirmations with updated (shorter) serial interval of COVID-19. The lesson learnt here is certainly useful in future similar outbreaks of COVID-19.

## Data Availability

We used publicly available data only.

## Declarations

### Ethics approval and consent to participate

The data were collected via public domain [5], and thus neither ethical approval nor individual consent was not applicable.

### Availability of materials

All data used in this work were publicly available via [5].

### Consent for publication

Not applicable.

## Acknowledgements

None.

## Disclaimer

The funding agencies had no role in the design and conduct of the study; collection, management, analysis, and interpretation of the data; preparation, review, or approval of the manuscript; or decision to submit the manuscript for publication.

## Authors’ Contributions

SZ and DH conceived the study, carried out the analysis, and drafted the first manuscript. All authors discussed the results, critically read and revised the manuscript, and gave final approval for publication.

## References

1. Huang C, Wang Y, Li X, Ren L, Zhao J, Hu Y, Zhang L, Fan G, Xu J, Gu X et al: Clinical features of patients infected with 2019 novel coronavirus in Wuhan, China. Lancet (London, England) 2020.

2. Li Q, Guan X, Wu P, Wang X, Zhou L, Tong Y, Ren R, Leung KSM, Lau EHY, Wong JY et al: Early Transmission Dynamics in Wuhan, China, of Novel Coronavirus–Infected Pneumonia. New England Journal of Medicine 2020.

3. Wu JT, Leung K, Leung GM: Nowcasting and forecasting the potential domestic and international spread of the 2019-nCoV outbreak originating in Wuhan, China: a modelling study. The Lancet 2020.

4. Statement on the second meeting of the International Health Regulations Emergency Committee regarding the outbreak of novel coronavirus (2019-nCoV), World Health Organization (WHO). [https://www.who.int/news-room/detail/30-01-2020-statement-on-the-second-meeting-of-the-international-health-regulations-(2005)-emergency-committee-regarding-the-outbreak-of-novel-coronavirus-(2019-ncov)]

5. Identification of novel coronavirus infection on cruise ship in quarantine at yokohama port. Ministry of Health, Labour and welfare of Japan. [https://www.mhlw.go.jp/stf/newpage_09425.html]

6. Althaus CL: Ebola superspreading. The Lancet Infectious diseases 2015, 15(5):507–508.

7. Riou J, Althaus CL: Pattern of early human-to-human transmission of Wuhan 2019 novel coronavirus (2019-nCoV), December 2019 to January 2020. Eurosurveillance 2020, 25(4).

8. You C, Deng Y, Hu W, Sun J, Lin Q, Zhou F, Pang CH, Zhang Y, Chen Z, Zhou X-H: Estimation of the Time-Varying Reproduction Number of COVID-19 Outbreak in China. medRxiv 2020:2020.2002.2008.20021253.

9. Nishiura H, Linton NM, Akhmetzhanov AR: Serial interval of novel coronavirus (2019-nCoV) infections. medRxiv 2020:2020.2002.2003.20019497.

10. Fan J, Huang T: Profile likelihood inferences on semiparametric varying-coefficient partially linear models. Bernoulli 2005, 11(6):1031–1057.

11. Zhao S, Lin Q, Ran J, Musa SS, Yang G, Wang W, Lou Y, Gao D, Yang L, He D et al: Preliminary estimation of the basic reproduction number of novel coronavirus (2019-nCoV) in China, from 2019 to 2020: A data-driven analysis in the early phase of the outbreak. International Journal of Infectious Diseases 2020.

12. Lipsitch M, Cohen T, Cooper B, Robins JM, Ma S, James L, Gopalakrishna G, Chew SK, Tan CC, Samore MH: Transmission dynamics and control of severe acute respiratory syndrome. Science 2003, 300(5627):1966–1970.

13. Lloyd-Smith JO, Schreiber SJ, Kopp PE, Getz WM: Superspreading and the effect of individual variation on disease emergence. Nature 2005, 438(7066):355–359.

14. Hartfield M, Alizon S. Introducing the outbreak threshold in epidemiology. PLoS pathogens. 2013 Jun;9(6).

15. Guan W-j, Ni Z-y, Hu Y, Liang W-h, Ou C-q, He J-x, Liu L, Shan H, Lei C-l, Hui DSC et al: Clinical characteristics of 2019 novel coronavirus infection in China. medRxiv 2020:2020.2002.2006.20020974.

16. Tuite AR, Fisman DN: Reporting, Epidemic Growth, and Reproduction Numbers for the 2019 Novel Coronavirus (2019-nCoV) Epidemic. Annals of Internal Medicine 2020.

